# Retinopathy is more common, but age-related macular degeneration carries more visual impairment: a cross-sectional study of US adults graded from the same fundus photographs

**DOI:** 10.64898/2026.11.08.26367574

**Authors:** Yijie Tang, Ruijie Guo, Hao Cheng

## Abstract

**Background:** Retinopathy and age-related macular degeneration (AMD) are the most common findings on fundus photographs of older adults, yet their frequency, correlates and visual consequences have been estimated in separate samples. We compared them in a national survey that graded both from the same images.

**Methods:** We analyzed 5593 adults aged ≥40 years in the US National Health and Nutrition Examination Survey 2005–2008 with photographs gradable for both conditions. We estimated survey-weighted prevalence, fitted identically specified logistic models for retinopathy (participants with diabetes) and AMD (full sample), and compared presenting visual impairment (better-eye acuity worse than 20/40) and self-rated vision across four groups: neither condition, retinopathy only, AMD only, and both.

**Results:** Retinopathy was more prevalent than AMD (9.54% [95% confidence interval (CI) 8.52–10.56] vs 6.52% [5.54–7.50]); AMD overtook it only from age 70 years. In participants with diabetes, retinopathy was associated with HbA1c (odds ratio [OR] 1.54 per percentage point; 1.33–1.78) and male sex (1.54; 1.10–2.17). AMD was associated with age (OR 1.09 per year; 1.07–1.11) and was less common in non-Hispanic Black adults (0.48; 0.33–0.71). Presenting impairment affected 4.91% of participants with neither condition, 7.09% with retinopathy only, 14.57% with AMD only and 26.15% with both. After age adjustment, impairment remained associated with AMD only (OR 2.23; 1.54–3.23) and both conditions (4.09; 2.28–7.37) but not retinopathy only (1.32; 0.91–1.90). Neither retinopathy nor diabetes was associated with AMD. Self-rated vision agreed poorly with measured impairment (κ = 0.15): only 12% of impaired participants rated their vision poor or very poor.

**Conclusions:** Retinopathy is the more frequent retinal finding in US adults aged ≥40 years, but AMD carries more of the measured visual impairment, independently of age. Grading diabetic screening photographs for AMD, and measuring acuity rather than asking about vision, would reach more people who see poorly.

## Background

Retinopathy and age-related macular degeneration (AMD) are among the most common findings when the retina of an older adult is photographed, and both are leading retinal causes of visual impairment in ageing populations [1–6]. Beyond that, they have little in common.

Retinopathy is a disorder of the retinal microvasculature, ranging from an isolated microaneurysm or hemorrhage to proliferative disease, and it is studied mostly in people with diabetes, in whom chronic hyperglycemia is its main driver [1, 2]. AMD is a degenerative disease of the macula in which drusen and pigmentary change may progress to geographic atrophy or neovascularization, and its dominant correlate is age [3, 7–10].

Because both conditions concentrate in older adults and appear on the same fundus photograph, services that plan eye care or retinal screening for an ageing population have to deal with both at once. The evidence, however, has accumulated separately. Different samples, grading protocols and definitions of visual loss make it hard to say, for a single population, which condition is more common, who is affected by each, and how much vision each costs.

The US National Health and Nutrition Examination Survey (NHANES) 2005–2008 can answer these questions. It remains the most recent nationally representative US survey in which adults aged 40 years and older underwent fundus photography graded for both retinopathy and AMD, with visual acuity measured at the same examination [11–13]. The landmark analyses of these data each addressed one question. Zhang et al. found retinopathy in 28.5% (95% confidence interval [CI] 24.9–32.5) of adults with diabetes [11]. Klein et al. reported any AMD in 6.5% (5.5–7.6) and late AMD in 0.8% (0.5–1.3) of adults aged 40 years and older [12]. Chou et al. showed that presenting visual impairment affected 7.5% (6.9–8.1) of the same age group, about two-thirds of it correctable with refraction, and that AMD was strongly associated with nonrefractive impairment (odds ratio [OR] 4.52; 95% CI 2.50–8.17) whereas retinopathy was not (OR 2.55; 0.68–9.51) [13]. Elsewhere, the relation between diabetes or diabetic retinopathy and AMD has been examined in population cohorts, claims data and a meta-analysis [14–17], and cardiovascular correlates of AMD in the Beaver Dam Eye Study [18].

Those reports used different analytic samples (1006, 5553 and 5222 participants) and different models, so their estimates cannot be set side by side. We therefore re-analyzed NHANES 2005–2008 in one fixed sample of adults whose photographs were gradable for both conditions, applying the same methods to each. We asked four questions. How common is each condition, overall and across age, sex and race/ethnicity? Which factors are independently associated with each when an identical model is used? How do measured visual acuity and self-rated vision differ between people with neither condition, one condition or both? And are the two conditions associated with each other?

## Methods

### Data source and participants

NHANES is a continuous program of cross-sectional surveys of the civilian, noninstitutionalized US population that combines a household interview with a standardized examination in a mobile examination center (MEC) [19]. We pooled the 2005–2006 and 2007–2008 cycles and linked nine public-use modules per cycle (demographics, retinal imaging, vision examination, vision questionnaire, diabetes questionnaire, glycohemoglobin, blood pressure, body measures and smoking) by participant identifier. Coding was identical across cycles except for age, which was top-coded at 85 years in 2005–2006 and at 80 years in 2007–2008; we capped age at 80 years in both cycles. Source files, download dates and checksums are archived with the analysis code [20], and the NHANES variables used are listed in Additional file 1: Table S1.

Participants were eligible if they were aged 40 years or older, attended the MEC examination and had a person-level (worse-eye) grade for both retinopathy and AMD. All eligible participants were included. The study is reported according to the STROBE statement [21].

### Fundus photography and grading

Two 45° non-mydriatic digital photographs were taken of each eye, one centered on the macula and one on the optic disc, with a Canon CR6-45NM camera fitted with an EOS 10D digital back. Images were graded at the University of Wisconsin reading center by graders masked to diabetes status, and every photographed participant was graded for both conditions whether or not they had diabetes [11–13]. Retinopathy was graded with NHANES categories adapted from the Early Treatment Diabetic Retinopathy Study classification [22, 23], and AMD with a modified Wisconsin Age-Related Maculopathy Grading System [24]. The NHANES files label AMD as “age-related maculopathy”; we use AMD throughout.

### Definitions

Retinopathy was defined as a worse-eye retinopathy level of 2 or higher, which corresponds to mild retinopathy or worse [23]. Because grading did not depend on diabetes status, retinopathy in the full sample includes participants without diabetes; we use the term diabetic retinopathy only for retinopathy within the diabetes subpopulation. AMD was defined as early or late AMD in the worse eye, comparable to the “any AMD” definition of Klein et al. [12]. Late AMD (53 participants) was too infrequent to analyze separately. From the two outcomes we formed four mutually exclusive groups: neither condition, retinopathy only, AMD only, and both.

Diabetes was defined as self-reported diagnosed diabetes (a “borderline” response was not counted) or an HbA1c of 6.5% or higher, as in Zhang et al. [11]. Diabetes defined the population for the retinopathy model but did not restrict the prevalence estimates. Among participants with self-reported diabetes, duration was calculated as age minus self-reported age at diagnosis, and insulin use as current use reported at interview.

Visual acuity was measured as presenting acuity, with habitual distance correction if any, using a Nidek ARK-760 autorefractor; eyes with presenting acuity of 20/30 or worse were retested after objective refraction [19]. Subjectively refined best-corrected acuity was not measured in these cycles [13]. We used the better eye, or the only eye measured. Visual impairment was defined as better-eye presenting acuity worse than 20/40, and refraction-aided impairment, a secondary outcome, as better-eye acuity still worse than 20/40 after objective refraction. For continuous analysis, acuity was converted to logMAR; the NHANES category “worse than 20/200” (4 participants) was scored as 20/200 (logMAR 1.0). Self-rated vision came from the vision questionnaire (five categories, excellent to very poor) and was dichotomized as poor or very poor versus better; “don’t know” was treated as missing.

Both regression models included the same covariates: age (years), sex, race/ethnicity (non-Hispanic White, non-Hispanic Black, Mexican American, other Hispanic, and other or multiracial), HbA1c (%), body mass index (kg/m²), systolic blood pressure (mean of the first three readings, mmHg) and lifetime smoking of at least 100 cigarettes, giving ten parameters per model.

### Statistical analysis

All estimates accounted for the complex survey design, using the MEC examination weights divided by two for the pooled cycles, 31 strata and 62 primary sampling units (31 design degrees of freedom), with variances estimated by Taylor series linearization [25, 26].

Subpopulations were analyzed as domains rather than by subsetting the data, so that all participants contributed to variance estimation. Tests were two-sided at α = 0.05. Counts, group sizes and baseline characteristics are unweighted; prevalence, odds ratios, proportions with visual impairment and mean logMAR are survey-weighted.

Weighted prevalence of retinopathy and AMD was estimated with 95% Wald CIs based on the t distribution, overall and by age group (40–49, 50–59, 60–69 and 70 years or older), sex and race/ethnicity. Because the race/ethnicity groups differ in age structure, we also standardized their prevalence directly to the weighted age distribution of the analytic sample in nine bands (40–44 to 75–79 years, and 80 years), with CIs for the standardized estimates obtained by Taylor linearization.

To examine associated factors we fitted two survey-weighted logistic regression models with the same specification. The retinopathy model was fitted in the diabetes subpopulation, the conventional setting for diabetic retinopathy [11, 13], and the AMD model in the full sample. Retinopathy therefore appears in two forms in this paper, as any retinopathy in the full sample for prevalence and as diabetic retinopathy for modeling, and we state which applies throughout. Because HbA1c contributes to the definition of diabetes, we refitted the retinopathy model in participants with self-reported diabetes, whose diabetes status does not depend on HbA1c, adding diabetes duration and current insulin use. We also fitted the AMD model with retinopathy, and separately with diabetes, as the exposure of interest, keeping the same covariates.

For vision, we estimated the weighted proportion with presenting visual impairment, and secondarily refraction-aided impairment, in each of the four groups, together with the weighted mean better-eye logMAR. Differences in logMAR between groups were tested with an overall Wald F test from survey-weighted linear regression. Because the groups differ markedly in age, we compared presenting impairment across them in a survey-weighted logistic model adjusted for age, and estimated age-adjusted mean logMAR at the sample mean age. Agreement between measured impairment and self-rated poor or very poor vision was summarized at the individual level by unweighted Cohen’s κ.

Eight sensitivity analyses were specified in advance (Table 4): a stricter retinopathy threshold of level 3 or higher (S1); the four groups redefined using diabetic retinopathy only (S2); eye-specific rather than person-level grades (S3); restriction to participants with both eyes gradable (S4); comparison of participants with and without an HbA1c measurement (S5); alternative visual acuity thresholds (S6); the retinopathy model refitted in the full sample, giving a same-sample comparison with the AMD model (S7); and separate estimates for each survey cycle (S8). S8 also served as a check on the 2007–2008 glycohemoglobin file, which NCHS withdrew in November 2011 and republished unchanged in March 2012. We used 10 events per model parameter as a guide for fitting models [27, 28].

Covariates were handled by complete-case analysis. Within the analytic sample, HbA1c was missing for 177 participants (3.16%), systolic blood pressure for 104 (1.86%), body mass index for 43 (0.77%) and smoking status for 2 (0.04%) (Additional file 1: Table S2), leaving 984 participants (297 events) for the retinopathy model and 5287 (409 events) for the AMD model. The 34 participants with no acuity measurement in either eye were excluded from the vision analyses (n = 5559), and 18 participants with presenting impairment but no refraction-aided measurement were excluded from the secondary outcome.

The statistical analysis plan was finalized on 7 October 2026, after preliminary data checks and before the main analyses. Analyses added after the plan were the age-adjusted comparison of visual impairment, refraction-aided impairment by group, the AMD models with retinopathy or diabetes as exposure, the agreement analysis, a three-category coding of smoking within S7, the self-reported diabetes model and age-standardized prevalence.

Analyses were performed in SAS/STAT 9.4 (TS1M8) on SAS OnDemand for Academics; the self-reported diabetes model and age standardization were performed in R 4.5.2 with the survey package, version 4.5 [29]. Every reported value was checked against independently computed reference values. The analysis code and derived dataset are publicly available [20]. Large language model tools were used to assist with language editing, reference formatting, and figure preparation. All analyses were designed, performed, and verified by the authors, who reviewed all AI-assisted output and take full responsibility for the integrity and accuracy of this work.

## Results

### Study sample

Of 7081 participants aged 40 years or older, 6797 attended the MEC examination and 5828 underwent fundus photography; 5593 had photographs gradable for both conditions and formed the analytic sample (Fig. 1). Of these, 4542 had neither condition, 616 retinopathy only, 378 AMD only and 57 both. Participants with AMD were older than the rest, with mean ages of 69.6 years in the AMD-only group and 72.5 years in the group with both conditions, compared with 58.1 years in the group with neither. The retinopathy-only group had the highest mean HbA1c (6.75%) and among the highest systolic blood pressures (Table 1).

**Fig. 1.**
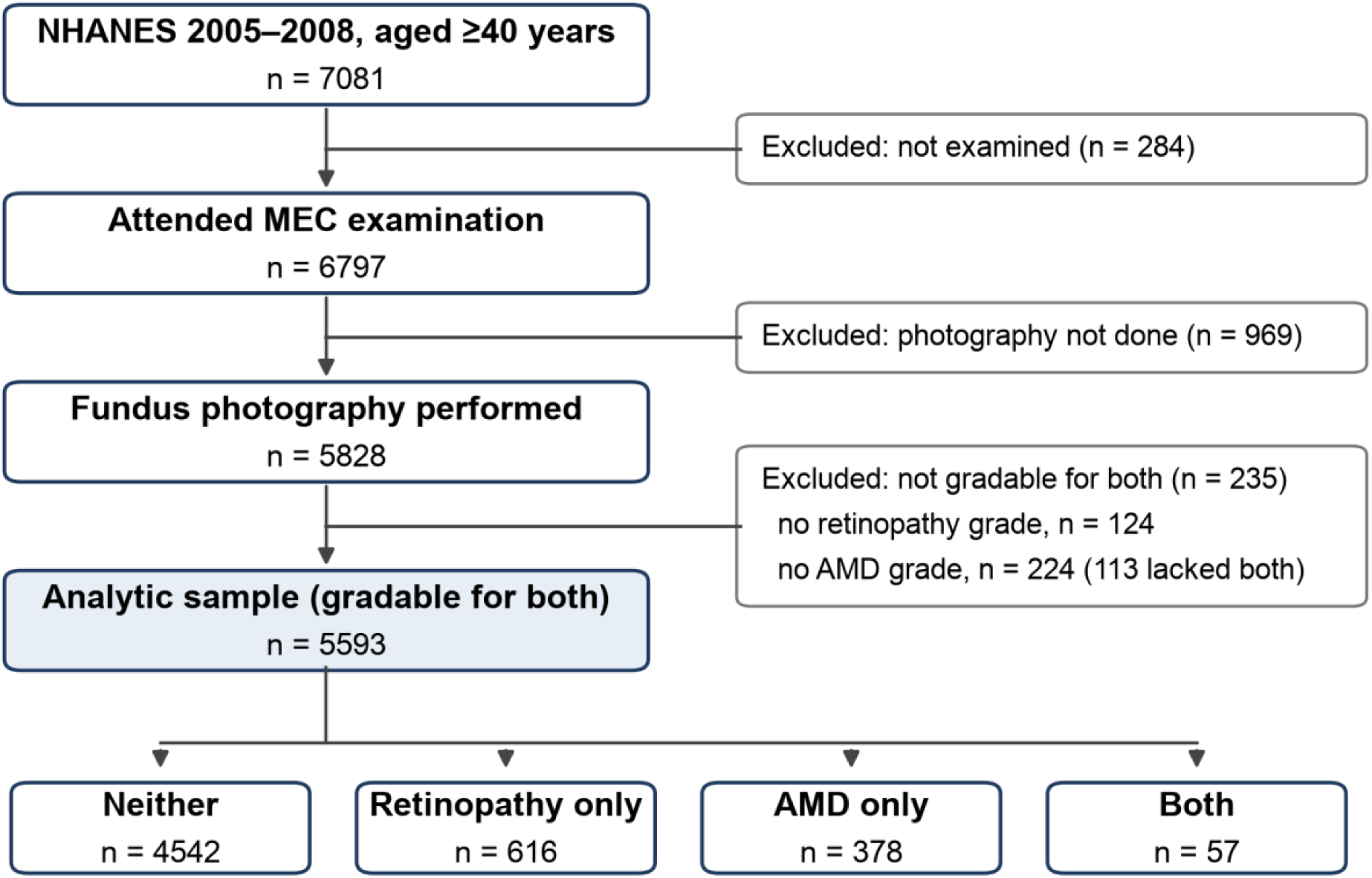
Selection of the analytic sample, NHANES 2005–2008. Counts are unweighted. Of the 5828 participants photographed, 124 lacked a retinopathy grade and 224 an AMD grade; 113 lacked both. AMD, age-related macular degeneration; MEC, mobile examination center

**Table 1.** Characteristics of participants by retinopathy and AMD status (unweighted)

| **Characteristic** | **Neither (n = 4542)** | **Retinopathy only (n = 616)** | **AMD only (n = 378)** | **Both (n = 57)** |
|---|---|---|---|---|
| Age, years, mean (SD) | 58.08 (11.92) | 61.03 (11.41) | 69.57 (11.07) | 72.53 (8.47) |
| Male, % | 49.16 | 56.33 | 52.38 | 49.12 |
| Non-Hispanic White, % | 53.92 | 40.58 | 71.69 | 66.67 |
| Body mass index, kg/m², mean (SD) | 29.17 (6.55) | 30.14 (6.43) | 28.34 (5.59) | 29.27 (5.60) |
| HbA1c, %, mean (SD) | 5.72 (0.90) | 6.75 (1.79) | 5.75 (0.76) | 6.15 (1.27) |
| Systolic blood pressure, mmHg, mean (SD) | 127.32 (18.95) | 135.43 (23.19) | 132.87 (22.19) | 137.91 (25.21) |
| Ever smoker (≥100 cigarettes), % | 52.56 | 52.11 | 55.82 | 54.39 |
| Better-eye presenting VA, Snellen denominator, mean (SD) | 26.40 (20.43) | 28.22 (30.34) | 34.10 (41.87) | 42.28 (40.85) |
| Visual impairment (better eye worse than 20/40), % | 7.13 | 9.30 | 16.62 | 26.32 |

Values are means (SD) or percentages of participants with non-missing data. Denominators for visual impairment are participants with acuity measured in at least one eye (4516, 613, 373 and 57). A larger Snellen denominator indicates worse acuity. AMD, age-related macular degeneration; HbA1c, glycated hemoglobin; SD, standard deviation; VA, visual acuity

Diabetes was present in 1050 participants: 830 by self-report and 220 by HbA1c alone. Just over half of the participants with retinopathy (352 of 673, unweighted) did not meet the definition of diabetes, and almost all of them (340 of 352) had mild retinopathy (level 2).

### Prevalence

The weighted prevalence of retinopathy was 9.54% (95% CI 8.52–10.56) and that of AMD 6.52% (5.54–7.50) (Table 2). Among participants with diabetes, 27.83% (24.21–31.44) had retinopathy. Both conditions became more common with age, but at very different rates (Fig. 2). Retinopathy roughly doubled between ages 40–49 and 70 years or older (6.17% to 13.46%), whereas AMD increased almost ninefold (2.30% to 20.23%) and was the more common condition only in the oldest group. Retinopathy was more common in men than in women (11.07% vs 8.16%); AMD was not (6.48% vs 6.56%). By race/ethnicity the two conditions ran in opposite directions: retinopathy was most prevalent in non-Hispanic Black participants (15.16%) and least prevalent in non-Hispanic White participants (8.42%), while AMD was most prevalent in non-Hispanic White participants (7.32%) and least prevalent in non-Hispanic Black participants (2.59%).

**Fig. 2.**
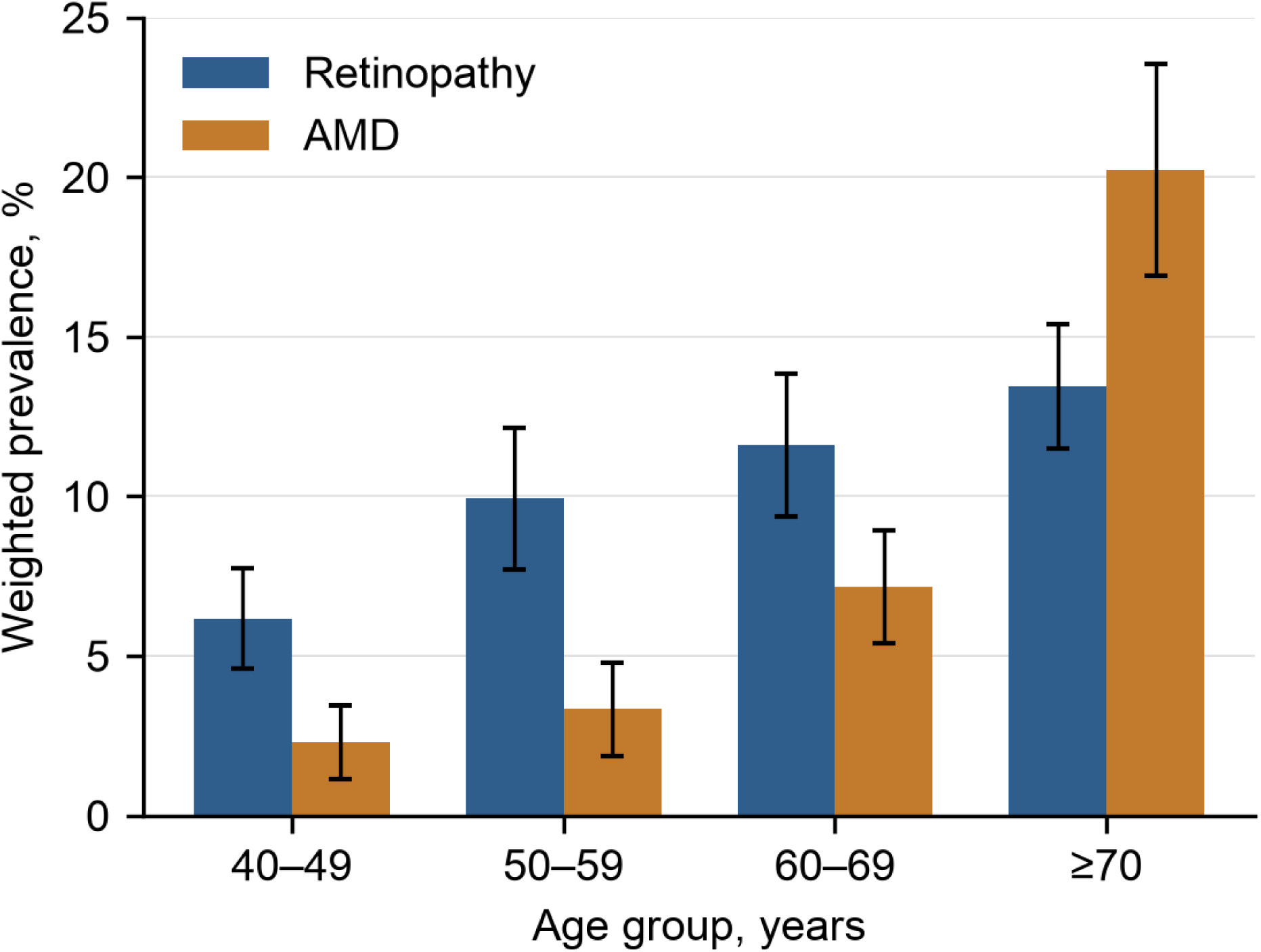
Weighted prevalence of retinopathy and AMD by age group. Bars show survey-weighted prevalence and error bars 95% confidence intervals (n = 5593). Retinopathy is any retinopathy regardless of diabetes status. AMD, age-related macular degeneration

**Table 2.** Weighted prevalence of retinopathy and AMD, overall and by subgroup.

| Subgroup | n | Retinopathy, % (95% CI) | AMD, % (95% CI) |
|---|---|---|---|
| **Overall** | **5593** | **9.54 (8.52–10.56)** | **6.52 (5.54–7.50)** |
| *Age, years* |  |  |  |
| 40–49 | 1501 | 6.17 (4.61–7.73) | 2.30 (1.14–3.45) |
| 50–59 | 1327 | 9.94 (7.72–12.15) | 3.34 (1.87–4.80) |
| 60–69 | 1386 | 11.61 (9.38–13.84) | 7.18 (5.40–8.95) |
| ≥70 | 1379 | 13.46 (11.50–15.41) | 20.23 (16.91–23.54) |
| *Sex* |  |  |  |
| Men | 2806 | 11.07 (9.80–12.34) | 6.48 (5.29–7.67) |
| Women | 2787 | 8.16 (6.98–9.34) | 6.56 (5.46–7.66) |
| *Race/ethnicity* |  |  |  |
| Non-Hispanic White | 3008 | 8.42 (7.28–9.57) | 7.32 (6.09–8.55) |
| Non-Hispanic Black | 1138 | 15.16 (12.59–17.74) | 2.59 (1.70–3.48) |
| Mexican American | 864 | 12.59 (10.39–14.79) | 5.08 (3.62–6.53) |
| Other Hispanic<sup>a</sup> | 401 | 14.69 (10.37–19.00) | 4.40 (2.23–6.57) |
| Other/multiracial | 182 | 9.33 (3.93–14.73) | 4.51 (1.63–7.40) |

Survey-weighted estimates with 95% CIs from Taylor series linearization (31 design degrees of freedom); n is the unweighted number of participants. Retinopathy is any retinopathy (worse-eye level ≥2) regardless of diabetes status; AMD is early or late AMD. <sup>a</sup>NCHS does not recommend separate estimates for other Hispanic participants in these cycles [25]; they are shown for completeness. AMD, age-related macular degeneration; CI, confidence interval

Age standardization left the contrasts for non-Hispanic Black participants intact: their standardized prevalence was 15.75% (13.00–18.51) for retinopathy, against 8.22% (7.08–9.36) in non-Hispanic White participants, and 3.04% (2.04–4.03) for AMD, against 6.87% (5.92–7.81) (Additional file 1: Table S4). The lower crude AMD prevalence among Mexican American participants, by contrast, disappeared (standardized 6.73%; 4.97–8.49), as it did among other Hispanic participants (6.07%; 3.90–8.24).

### Factors associated with each condition

In the diabetes subpopulation (984 participants with complete data, 297 with retinopathy), retinopathy was associated with higher HbA1c (OR 1.54 per percentage point; 95% CI 1.33–1.78; p < 0.001), male sex (OR 1.54; 1.10–2.17; p = 0.015) and older age (OR 1.02 per year; 1.00–1.04; p = 0.038) (Table 3, Fig. 3). Ever-smoking was associated with lower odds of retinopathy (OR 0.64; 0.47–0.88; p = 0.007), and the association with systolic blood pressure was borderline (OR 1.01 per mmHg; 1.00–1.02; p = 0.053). Non-Hispanic Black participants had an adjusted OR of 1.41 (0.83–2.38) relative to non-Hispanic White participants.

**Fig. 3.**
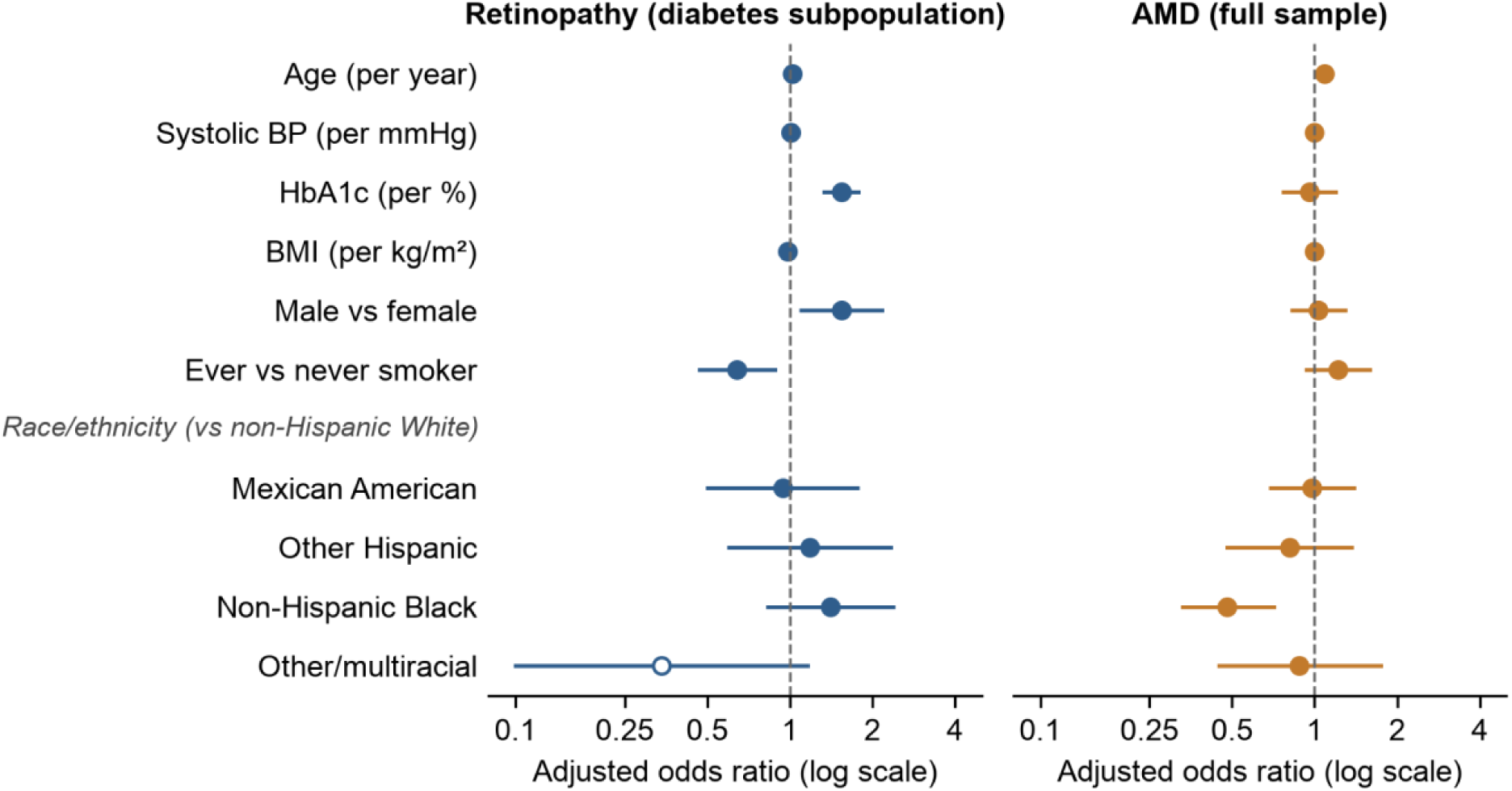
Adjusted odds ratios for retinopathy and AMD from identically specified models. Left, retinopathy in the diabetes subpopulation (984 complete cases, 297 events); right, AMD in the full sample (5287 complete cases, 409 events). Points are odds ratios and lines 95% confidence intervals on a log scale. Reference categories are female, non-Hispanic White and never smoker. The open marker denotes a sparse category (7 events). AMD, age-related macular degeneration; BMI, body mass index; BP, blood pressure

**Table 3.** Adjusted odds ratios for retinopathy (diabetes subpopulation) and AMD (full sample)

| **Variable** | **Retinopathy, OR (95% CI)** | **p** | **AMD, OR (95% CI)** | **p** |
|---|---|---|---|---|
| Age (per year) | 1.02 (1.00–1.04) | 0.038 | 1.09 (1.07–1.11) | <0.001 |
| Systolic blood pressure (per mmHg) | 1.01 (1.00–1.02) | 0.053 | 1.00 (0.99–1.01) | 0.997 |
| HbA1c (per percentage point) | 1.54 (1.33–1.78) | <0.001 | 0.96 (0.77–1.19) | 0.694 |
| Body mass index (per kg/m²) | 0.98 (0.95–1.01) | 0.248 | 1.00 (0.98–1.03) | 0.775 |
| Male vs female | 1.54 (1.10–2.17) | 0.015 | 1.03 (0.83–1.29) | 0.773 |
| Ever vs never smoker | 0.64 (0.47–0.88) | 0.007 | 1.22 (0.93–1.59) | 0.146 |
| *Race/ethnicity (vs non-Hispanic White)* |  |  |  |  |
| Mexican American | 0.94 (0.50–1.76) | 0.839 | 0.98 (0.69–1.39) | 0.904 |
| Other Hispanic | 1.18 (0.60–2.33) | 0.618 | 0.81 (0.48–1.36) | 0.410 |
| Non-Hispanic Black | 1.41 (0.83–2.38) | 0.193 | 0.48 (0.33–0.71) | <0.001 |
| Other/multiracial<sup>a</sup> | 0.34 (0.10–1.16) | 0.083 | 0.88 (0.45–1.74) | 0.712 |

Both models include all variables shown and are survey-weighted. Retinopathy model: diabetes subpopulation (n = 1050; 984 complete cases, 297 events). AMD model: full sample (n = 5593; 5287 complete cases, 409 events). Never smoker: fewer than 100 cigarettes in lifetime. p values are from Wald tests. <sup>a</sup>In the retinopathy model this estimate is based on 7 events among 28 participants. AMD, age-related macular degeneration; CI, confidence interval; HbA1c, glycated hemoglobin; OR, odds ratio

**Table 4.** Prespecified sensitivity analyses.

|  | Analysis | Result | Interpretation |
|---|---|---|---|
| S1 | Retinopathy threshold raised to level ≥3 | 110 participants with retinopathy (98 with diabetes); 88 events in complete cases (8.8 per parameter) | Model not fitted (fewer than 10 events per parameter) |
| S2 | Groups redefined using diabetic retinopathy only | Groups: 4861 / 297 / 411 / 24; retinopathy among participants with diabetes 27.83% (24.21–31.44) | Consistent with Zhang et al. [11] |
| S3 | Eye-specific grades | Retinopathy: right eye 460, left eye 425 (with diabetes 261, 236); AMD: right eye 333, left eye 296 | Person-level results not driven by one eye |
| S4 | Both eyes gradable | n = 4996; retinopathy 588, AMD 386; diabetes 889, of whom 271 had retinopathy | Unchanged |
| S5 | Participants with vs without HbA1c | HbA1c missing for 177 (3.16%), 38 of them in the diabetes subpopulation | Missingness limited |
| S6 | Visual acuity thresholds | Better-eye presenting acuity worse than 20/50: 124; worse than 20/60: 77; worse than 20/80 (20/200 or worse): 40; worse than 20/200: 4 | Group ordering unchanged |
| S7 | Retinopathy model refitted in full sample | n = 5593; 673 events, 623 in complete cases (62.3 per parameter) | Pattern matches the diabetes-subpopulation model, not the AMD model (Additional file 1: Table S3) |
| S8 | Each survey cycle separately (design df 15 and 16) | 2005–2006 (n = 2410): retinopathy 9.48% (8.32–10.65), AMD 6.50% (4.79–8.21). 2007–2008 (n = 3183): retinopathy 9.59% (7.83–11.35), AMD 6.54% (5.40–7.69). Mean HbA1c 5.75% (SD 1.03) and 5.90% (SD 1.11) | Stable across cycles |

Counts are unweighted; prevalence estimates are survey-weighted with 95% CIs. Groups in S2 are neither, diabetic retinopathy only, AMD only and both. AMD, age-related macular degeneration; CI, confidence interval; df, degrees of freedom; HbA1c, glycated hemoglobin; SD, standard deviation

Among participants with self-reported diabetes (759 with complete data, 262 with retinopathy), the association with HbA1c persisted (OR 1.44 per percentage point; 1.22–1.70; p < 0.001) after duration and insulin use were added (Additional file 1: Table S5).

Retinopathy was also associated with longer duration (OR 1.06 per year; 1.03–1.10; p < 0.001), current insulin use (3.40; 2.11–5.48; p < 0.001), male sex (1.89; 1.31–2.71; p = 0.001) and systolic blood pressure (1.01 per mmHg; 1.00–1.03; p = 0.008). With duration in the model, age was no longer associated with retinopathy (OR 1.01; 0.98–1.03).

In the full sample (5287 participants with complete data, 409 with AMD), AMD was associated with age (OR 1.09 per year; 1.07–1.11; p < 0.001), roughly a 2.4-fold increase in odds per decade, and was less common in non-Hispanic Black than in non-Hispanic White participants (OR 0.48; 0.33–0.71; p < 0.001). HbA1c, blood pressure, body mass index, sex and smoking were not associated with AMD. With the same covariates, neither retinopathy (OR 1.08; 0.70–1.67) nor diabetes (OR 0.97; 0.66–1.43) was associated with AMD.

The sensitivity analyses are summarized in Table 4. Raising the retinopathy threshold to level 3 or higher (S1) left 88 events in the complete-case diabetes subpopulation, 8.8 per parameter, and the model was not fitted. Restricting the groups to diabetic retinopathy (S2), using eye-specific grades (S3), restricting to participants with both eyes gradable (S4) and varying the visual acuity threshold (S6) did not change the pattern of results. Refitted in the full sample (S7), the retinopathy model kept its own pattern — higher HbA1c (OR 1.84 per percentage point; 95% CI 1.61–2.11), male sex (1.53; 1.25–1.88) and higher odds among non-Hispanic Black participants (1.47; 1.07–2.01) — rather than converging on the AMD pattern, in which age dominated and non-Hispanic Black participants had lower odds (Table 3; Additional file 1: Table S3). The correlates of the two conditions therefore differ even within one sample. Estimates from the two survey cycles were close: retinopathy 9.48% in 2005–2006 and 9.59% in 2007–2008, and AMD 6.50% and 6.54% (S8).

### Visual impairment

Overall, 5.82% (95% CI 5.07–6.58) of participants had presenting visual impairment. The proportion rose from 4.91% (4.23–5.60) among participants with neither condition to 7.09% (4.93–9.25) with retinopathy only, 14.57% (9.86–19.27) with AMD only and 26.15% (14.22–38.08; 15 of 57) with both (Fig. 4). Weighted mean better-eye logMAR followed the same order (0.069, 0.079, 0.134 and 0.207; Wald F(3, 31) = 18.8, p < 0.001).

**Fig. 4.**
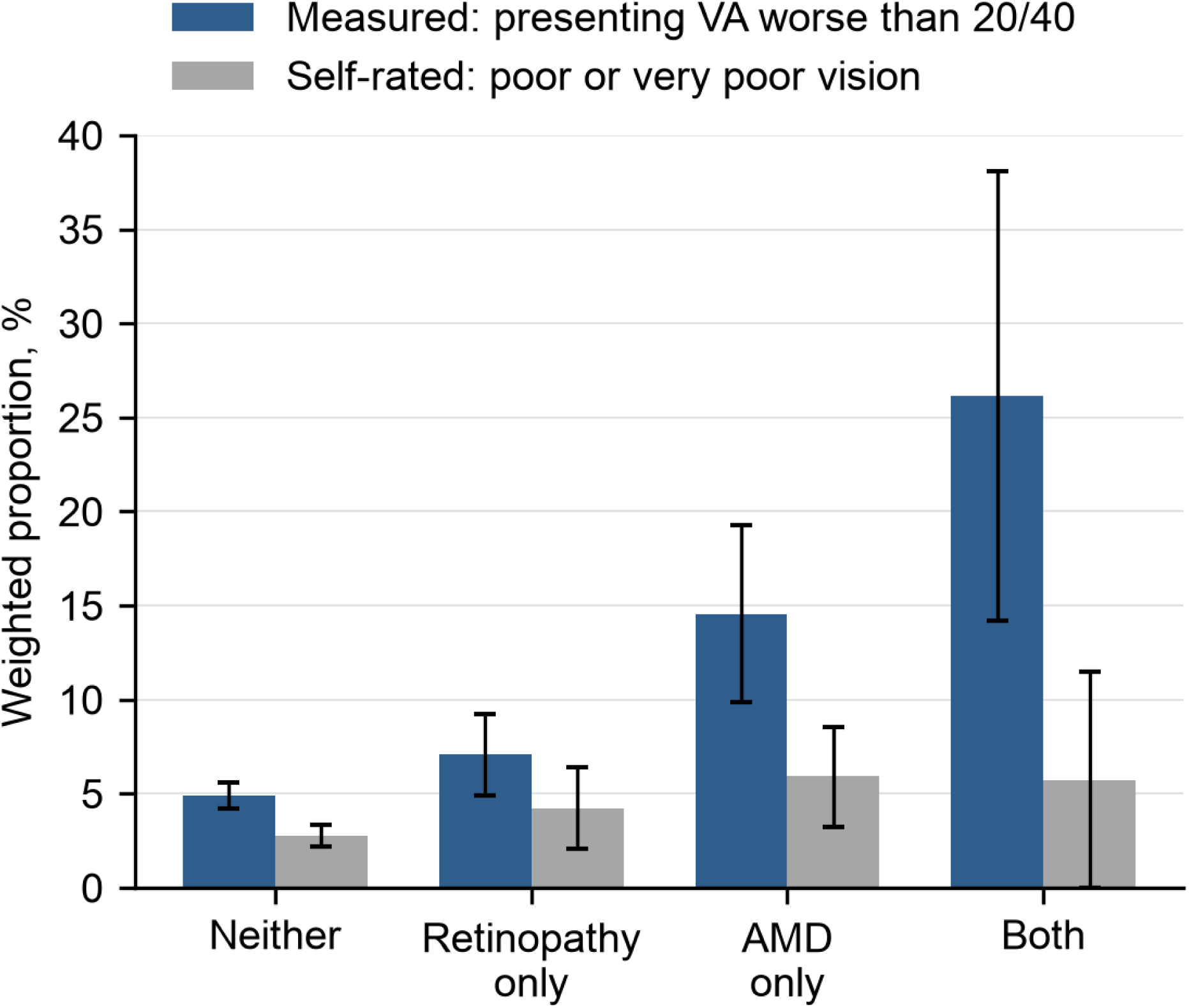
Measured visual impairment and self-rated poor vision by disease group. Bars show survey-weighted proportions and error bars 95% confidence intervals. Blue, better-eye presenting acuity worse than 20/40 (n = 5559); grey, self-rated poor or very poor vision (n = 5588). The lower confidence limit for self-rated vision in the group with both conditions (57 participants) is truncated at 0. AMD, age-related macular degeneration; VA, visual acuity

Adjustment for age separated the two conditions. Relative to participants with neither condition, the odds of impairment were not clearly raised with retinopathy only (OR 1.32; 0.91–1.90) but were more than doubled with AMD only (2.23; 1.54–3.23) and quadrupled with both conditions (4.09; 2.28–7.37). Age-adjusted mean logMAR at the sample mean age of 56.3 years was 0.071, 0.074, 0.110 and 0.172.

After objective refraction the gradient was steeper. Among 5541 participants with a refraction-aided measurement, 93 remained impaired: 0.53% (0.36–0.69) with neither condition, 1.65% (0.34–2.95) with retinopathy only, 5.53% (3.08–7.99) with AMD only and 10.69% (2.75–18.63) with both. Impairment was thus about ten times as common with AMD only as with neither condition, compared with three times for presenting acuity.

Most measured impairment was mild. Of the 456 participants with presenting impairment, 332 (73%) had a better-eye acuity of 20/50, one line below the threshold, and 40 had 20/200 or worse (Table 4, S6).

Self-rated vision barely separated the groups. Among the 5588 participants who rated their vision, poor or very poor vision was reported by 2.75% of those with neither condition, 4.21% with retinopathy only, 5.92% with AMD only and 5.71% with both, a spread of about 3 percentage points against 21 for measured impairment. Agreement between the two measures was slight (unweighted κ = 0.15): 12% of participants with measured impairment rated their vision as poor or very poor, and 39% as fair or worse.

## Discussion

Graded from the same photographs in one national sample, retinopathy and AMD behaved as two different problems. Retinopathy was about one and a half times as common, but AMD carried more of the measured visual loss: with age held constant, impairment was more than twice as likely with AMD alone and four times as likely with both conditions, whereas retinopathy alone carried no clear excess. The conditions had different correlates — glycemia, diabetes duration and male sex for retinopathy, age for AMD — opposite race/ethnicity gradients, and no association with each other. Self-rated vision captured little of this.

Our estimates reproduce the single-disease analyses of the same survey: 27.83% retinopathy among adults with diabetes against 28.5% [11], and 6.52% any AMD against 6.5% [12]. The contrasts between the conditions therefore reflect the conditions, not an unusual sample or definition. Presenting impairment (5.82%) was lower than the 7.5% reported by Chou et al. [13], most likely because we required photographs gradable for both conditions. Ungradable non-mydriatic images in older adults usually result from media opacity, small pupils or unsteady fixation, all of which also reduce acuity, so this requirement removed a disproportionate share of people who see poorly. Our estimates of visual impairment are, if anything, conservative.

In people with diabetes, retinopathy tracked glycemia: the odds rose by about half for each percentage point of HbA1c, in line with its central role in diabetic retinopathy [1, 2]. The association was not an artifact of HbA1c helping to define the subpopulation. Among participants with self-reported diabetes, for whom HbA1c plays no part in the definition, it held (OR 1.44), and diabetes duration, one of the strongest correlates of diabetic retinopathy [1], and insulin use, a marker of more advanced diabetes, entered with the expected strength. Once duration was included, age lost its association, so the age gradient in the main model reflects time with diabetes rather than ageing. Systolic blood pressure, borderline in the main model, was associated in this fuller model. Male sex was associated with retinopathy in every model we fitted (ORs 1.53 to 1.89).

Non-Hispanic Black adults had the highest prevalence of retinopathy and the lowest of AMD, and both contrasts survived age standardization. For retinopathy, the adjusted OR for non-Hispanic Black participants was similar in every model (1.32 to 1.47) and was statistically significant in the full sample, where precision was greatest (1.47; 1.07–2.01). An excess that persists after adjustment for HbA1c, blood pressure and body mass index points to factors these single measurements do not capture, such as cumulative glycemic exposure and access to diabetes and eye care. The lower prevalence of AMD in Black adults agrees with multiethnic and earlier NHANES analyses [10, 12]. The lower crude prevalence of AMD among Mexican American adults, in contrast, vanished after standardization (6.73% vs 6.87% in non-Hispanic White adults) and in the adjusted model (OR 0.98), and so reflects the younger age structure of this group rather than lower age-specific occurrence.

Half of the retinopathy in this population occurred in people without diabetes, and almost all of it was mild. In the full-sample model it was associated with blood pressure as well as HbA1c (Additional file 1: Table S3), matching an analysis of participants without diabetes in this survey, in which 98% of retinopathy was minimal to mild and its correlates were male sex, systolic blood pressure and HbA1c [30]. The full-sample prevalence is therefore a prevalence of retinal microvascular signs, not of diabetic retinopathy, which is why we modeled retinopathy within the diabetes subpopulation.

Ever-smoking was inversely associated with retinopathy in the diabetes model (OR 0.64), but the association weakened among participants with self-reported diabetes once duration and insulin use were included (0.69; 0.45–1.05) and was absent in the full sample (0.94; 0.71–1.24), where former and current smokers diverged (0.77 and 1.23). This pattern points to confounding and selective survival of smokers with less vascular disease rather than protection, and mirrors a meta-analysis in which smoking was associated with lower risk of diabetic retinopathy in type 2 diabetes and higher risk in type 1 diabetes [31].

AMD was associated with age and little else, its odds rising about 2.4-fold per decade. Smoking, the strongest established modifiable risk factor for AMD [8, 9], was not associated, which most likely reflects the measure available: lifetime smoking of at least 100 cigarettes merges long-term former smokers with current smokers and carries no information on dose. Neither diabetes nor retinopathy was associated with AMD in identically adjusted models.

The meta-analytic association between diabetes and AMD is strongest for late disease [17], of which this sample contained 53 cases; for early AMD, which made up most cases here, our data give no indication of shared metabolic determinants.

The visual consequences of the two conditions show why prevalence is a poor guide to burden. Chou et al. found AMD, but not retinopathy, associated with nonrefractive impairment [13]; comparing the same people with age held constant sharpens that finding. AMD alone doubled the odds of presenting impairment, and after objective refraction, which removes most of the impairment due to uncorrected refractive error, the AMD-only group had ten times the impairment prevalence of people with neither condition. The anatomy explains the difference: AMD involves the macula by definition, whereas retinopathy at the threshold used here is mostly mild nonproliferative disease, which seldom reduces central acuity. One in four participants with both conditions was impaired. Photographs taken for diabetic eye screening already include the macula; grading them for AMD as well would extend screening, without additional imaging, to the condition that carried more of the visual loss.

Self-rated vision barely registered these differences. Only one in eight participants with measured impairment described their vision as poor or very poor, and fewer than two in five as even fair. Most of this impairment was mild — 73% of those affected read 20/50 — but 20/50 falls short of the 20/40 better-eye standard for an unrestricted driving licence in all but three US states [32]. Gradual, bilateral loss is easily accommodated, and expectations of sight decline with age. Measuring acuity finds these people; asking them does not.

### Strengths and limitations

Both conditions were graded on the same photographs, centrally and masked to diabetes status, in a national probability sample analyzed with design-based methods and identical models; the analysis plan, code and derived data are public. The design is cross-sectional, so the associations carry no temporal order. The data were collected in 2005–2008, and diabetes care and anti-vascular endothelial growth factor (VEGF) treatment of neovascular AMD have changed since, but no later US national survey has graded retinal photographs. Requiring gradable photographs for both conditions excluded 1204 participants (969 not photographed and 235 with ungradable images), which likely biases the prevalence of visual impairment downward. Two 45° non-mydriatic fields miss peripheral lesions, and NHANES measured presenting and refraction-aided rather than best-corrected acuity. Smoking was recorded as lifetime exposure, age was capped at 80 years, and estimates for the group with both conditions and for other Hispanic participants, for whom NCHS does not recommend separate estimates [25], are imprecise.

## Conclusions

In US adults aged 40 years and older, retinopathy is the more common retinal finding, but AMD carries more of the measured visual impairment, independently of age. The two conditions have different correlates and are not associated with each other, and self-rated vision misses most measured loss. Eye-care planning for ageing populations should therefore weigh visual consequences, not prevalence alone: grading screening photographs for AMD and measuring acuity rather than asking about vision would reach more of the people who see poorly.

## Abbreviations

AMD: Age-related macular degeneration
CI: Confidence interval
HbA1c: Glycated hemoglobin
logMAR: Logarithm of the minimum angle of resolution
MEC: Mobile examination center
NCHS: National Center for Health Statistics
NHANES: National Health and Nutrition Examination Survey
OR: Odds ratio
STROBE: Strengthening the Reporting of Observational Studies in Epidemiology
VA: Visual acuity
VEGF: Vascular endothelial growth factor

## Declarations

### Ethics approval and consent to participate

NHANES 2005–2006 was approved by the NCHS Research Ethics Review Board (protocol #2005-06), and the approval was continued for 2007–2008 [33]. All NHANES participants provided written informed consent. This secondary analysis used only publicly available, de-identified data.

### Consent for publication

Not applicable.

### Availability of data and materials

The NHANES data analyzed in this study are publicly available from the National Center for Health Statistics (https://wwwn.cdc.gov/nchs/nhanes/) [19]. The analysis code and derived analytic dataset are available in the Zenodo repository, https://doi.org/10.5281/zenodo.24070599 [20].

## Authors’ contributions

YT: conceptualization, methodology, software, formal analysis, data curation, writing – original draft, writing – review and editing, supervision and project administration. RG: conceptualization, methodology, software, formal analysis, visualization, writing – original draft, and writing – review and editing. HC: data curation, investigation, and writing – review and editing. All authors read and approved the final manuscript.

## Data Availability

The NHANES data analyzed in this study are publicly available from the National Center for Health Statistics (https://www.cdc.gov/nchs/nhanes/). The original survey was approved by the NCHS Research Ethics Review Board. The analysis code and derived analytic dataset supporting the conclusions of this article are available in the Zenodo repository (https://doi.org/10.5281/zenodo.24070599).

https://wwwn.cdc.gov/nchs/nhanes/continuousnhanes/default.aspx?BeginYear=2005

https://wwwn.cdc.gov/nchs/nhanes/continuousnhanes/default.aspx?BeginYear=2007

## Acknowledgements

We thank Tianhao Xia, Peixu Yin and Chao Tang for their contributions to project planning, data discussion and preliminary analysis.

## Clinical trial number

Not applicable.

## Additional files

Additional file 1 (.docx): Tables S1–S5, covering variables, missing data and supplementary analyses.

