## Additional file 1: Supplementary tables for "Retinopathy is more common, but age-related macular degeneration carries more visual impairment: a cross-sectional study of US adults graded from the same fundus photographs"

Reference numbers refer to the reference list of the main article.

**Table S1 NHANES variables and derived definitions**

| **Concept** | **NHANES variable(s)** | **Definition used** |
| --- | --- | --- |
| Age | RIDAGEYR | Years; capped at 80 in both cycles |
| MEC examination | RIDSTATR | Examined (RIDSTATR = 2) |
| Retinopathy (worse eye) | OPDURL4 | Level ≥2 (mild or worse) |
| AMD (worse eye) | OPDUARM | 1 = early, 2 = late; any AMD = ≥1 |
| Self-reported diabetes | DIQ010 | 1 = yes; borderline not counted |
| Age at diabetes diagnosis | DID040 | Duration = capped age − DID040; 777/999 = missing |
| Insulin use | DIQ050 | 1 = currently taking insulin |
| HbA1c | LBXGH | %; diabetes if ≥6.5 |
| Self-rated vision | VIQ031 | Poor or very poor (4–5) vs better; 9 = missing |
| Smoking | SMQ020 | Ever smoked ≥100 cigarettes |
| Current smoking | SMQ040 | Every day or some days vs not at all (S7 three-category coding) |
| Systolic blood pressure | BPX module, readings 1–3 | Mean of available first three readings |
| Presenting and refraction-aided acuity | VIX module | Better eye; Snellen denominator converted to logMAR; code 666 (worse than 20/200) scored as 20/200 |
| Survey design | SDMVSTRA, SDMVPSU, WTMEC2YR | 31 strata, 62 PSUs; weight = WTMEC2YR/2 |

AMD, age-related macular degeneration; HbA1c, glycated hemoglobin; MEC, mobile examination center; PSU, primary sampling unit

**Table S2 Missing data by variable and disease group**

| **Variable** | **Overall (n = 5593)** | **Neither (n = 4542)** | **Retinopathy only (n = 616)** | **AMD only (n = 378)** | **Both (n = 57)** |
| --- | --- | --- | --- | --- | --- |
| Age, sex, race/ethnicity | 0 (0.00) | 0 (0.00) | 0 (0.00) | 0 (0.00) | 0 (0.00) |
| HbA1c | 177 (3.16) | 140 (3.08) | 25 (4.06) | 9 (2.38) | 3 (5.26) |
| Systolic blood pressure | 104 (1.86) | 70 (1.54) | 22 (3.57) | 9 (2.38) | 3 (5.26) |
| Body mass index | 43 (0.77) | 38 (0.84) | 2 (0.32) | 2 (0.53) | 1 (1.75) |
| Smoking | 2 (0.04) | 2 (0.04) | 0 (0.00) | 0 (0.00) | 0 (0.00) |
| Better-eye visual acuity | 34 (0.61) | 26 (0.57) | 3 (0.49) | 5 (1.32) | 0 (0.00) |
| Self-rated vision | 5 (0.09) | 3 (0.07) | 2 (0.32) | 0 (0.00) | 0 (0.00) |
| Retinopathy and AMD grades | 0 (0.00) | 0 (0.00) | 0 (0.00) | 0 (0.00) | 0 (0.00) |

Values are unweighted numbers missing (%). AMD, age-related macular degeneration; HbA1c, glycated hemoglobin

**Table S3 Full-sample refit of the retinopathy model (S7)**

| **Variable** | **OR (95% CI)** | **p** |
| --- | --- | --- |
| Age (per year) | 1.02 (1.01–1.03) | 0.007 |
| Systolic blood pressure (per mmHg) | 1.01 (1.01–1.02) | <0.001 |
| HbA1c (per percentage point) | 1.84 (1.61–2.11) | <0.001 |
| Body mass index (per kg/m²) | 1.00 (0.98–1.01) | 0.520 |
| Male vs female | 1.53 (1.25–1.88) | <0.001 |
| Mexican American | 1.23 (0.87–1.75) | 0.230 |
| Other Hispanic | 1.58 (0.97–2.57) | 0.068 |
| Non-Hispanic Black | 1.47 (1.07–2.01) | 0.018 |
| Other/multiracial | 1.03 (0.51–2.07) | 0.931 |
| Ever vs never smoker | 0.94 (0.71–1.24) | 0.645 |

Full-sample refit of the retinopathy model (S7, Table 4): n = 5287 complete cases, 623 events; covariates identical to Table 3; survey-weighted (31 design df). In a three-category coding of smoking in this model, former smokers had OR 0.77 (0.56–1.07) and current smokers 1.23 (0.94–1.63) versus never smokers; SMQ040 was merged from the source NHANES files for this coding and will be added to the archived dataset in repository version 1.2.

**Table S4 Crude and age-standardized prevalence of retinopathy and AMD by race/ethnicity**

| **Race/ethnicity** | **n** | **Crude, % (95% CI)** | **Age-standardized, % (95% CI)** |
| --- | --- | --- | --- |
| *Retinopathy* |  |  |  |
| Non-Hispanic White | 3008 | 8.42 (7.28–9.57) | 8.22 (7.08–9.36) |
| Non-Hispanic Black | 1138 | 15.16 (12.59–17.74) | 15.75 (13.00–18.51) |
| Mexican American | 864 | 12.59 (10.39–14.79) | 13.70 (11.15–16.25) |
| Other Hispanicᵃ | 401 | 14.69 (10.37–19.00) | 14.59 (9.95–19.22) |
| Other/multiracial | 182 | 9.33 (3.93–14.73) | 8.61 (3.67–13.55) |
| *AMD* |  |  |  |
| Non-Hispanic White | 3008 | 7.32 (6.09–8.55) | 6.87 (5.92–7.81) |
| Non-Hispanic Black | 1138 | 2.59 (1.70–3.48) | 3.04 (2.04–4.03) |
| Mexican American | 864 | 5.08 (3.62–6.53) | 6.73 (4.97–8.49) |
| Other Hispanicᵃ | 401 | 4.40 (2.23–6.57) | 6.07 (3.90–8.24) |
| Other/multiracial | 182 | 4.51 (1.63–7.40) | 4.54 (1.91–7.17) |

Survey-weighted estimates (31 design degrees of freedom). Prevalence was directly standardized to the weighted age distribution of the analytic sample (n = 5593) in nine bands (40–44, 45–49, 50–54, 55–59, 60–64, 65–69, 70–74, 75–79 and 80 years); CIs for standardized estimates are from Taylor linearization of the standardized linear combination. Crude estimates are those in Table 2. ᵃNCHS does not recommend separate estimates for other Hispanic participants in these cycles [25]. AMD, age-related macular degeneration; CI, confidence interval

**Table S5 Retinopathy model in participants with self-reported diabetes, with diabetes duration and insulin use**

| **Variable** | **OR (95% CI)** | **p** |
| --- | --- | --- |
| Age (per year) | 1.01 (0.98–1.03) | 0.529 |
| Systolic blood pressure (per mmHg) | 1.01 (1.00–1.03) | 0.008 |
| HbA1c (per percentage point) | 1.44 (1.22–1.70) | <0.001 |
| Body mass index (per kg/m²) | 0.98 (0.95–1.01) | 0.157 |
| Male vs female | 1.89 (1.31–2.71) | 0.001 |
| Ever vs never smoker | 0.69 (0.45–1.05) | 0.083 |
| Diabetes duration (per year) | 1.06 (1.03–1.10) | <0.001 |
| Current insulin use vs none | 3.40 (2.11–5.48) | <0.001 |
| *Race/ethnicity (vs non-Hispanic White)* |  |  |
| Mexican American | 0.99 (0.49–2.03) | 0.988 |
| Other Hispanic | 1.22 (0.54–2.78) | 0.619 |
| Non-Hispanic Black | 1.32 (0.66–2.63) | 0.424 |
| Other/multiracial | 0.48 (0.16–1.38) | 0.166 |

Participants with self-reported diagnosed diabetes (n = 830; 759 complete cases, 262 events). Covariates as in Table 3, plus diabetes duration (age minus self-reported age at diagnosis) and current insulin use; survey-weighted logistic regression (31 design degrees of freedom); p values are from Wald tests. Never smoker: fewer than 100 cigarettes in lifetime. CI, confidence interval; HbA1c, glycated hemoglobin; OR, odds ratio
